# Multimodal Recruitment for an Internet-Based Pilot Study of Ovulation and Menstruation (OM) Health

**DOI:** 10.1101/2020.06.29.20142778

**Authors:** Shruthi Mahalingaiah, J. Jojo Cheng, Michael Winter, Erika Rodriguez, Victoria Fruh, Anna Williams, MyMy Nguyen, Rashmi Madhavan, Pascaline Karanja, Jill McCrae, Sai Charan Konanki, Kevin J Lane, Ann Aschengrau

**Affiliations:** Department of Obstetrics and Gynecology, Boston University School of Medicine, 85 East Concord Street, Boston, MA 02118, USA; Department of Epidemiology, Boston University School of Public Health, Talbot 3E, 715 Albany Street, Boston, MA 02118, USA; Department of Environmental Health, Harvard T.H. Chan School of Public Health, Boston, MA 02115, USA; Department of Biostatistics and Medical Informatics, University of Wisconsin, Madison, Wisconsin 53792, U.S.A.; Biostatistics and Epidemiology Data Analytics Center, Boston University School of Public Health, Fuller M-921B, 85 East Newton Street, Boston, MA 02118, USA; Department of Environmental Health, Boston University School of Public Health, Talbot, 715 Albany Street, Boston, MA 02118, USA

**Keywords:** Polycystic ovary syndrome (PCOS), menstrual cycle, multimodal recruitment strategy, epidemiology

## Abstract

**Background:** Multimodal recruitment strategies are a novel way to increase diversity of research populations. However, these methods have not been previously applied to understanding the prevalence of menstrual disorders such as polycystic ovary syndrome (PCOS).

**Methods:** We conducted the Ovulation and Menstruation Health (OM) Pilot Study using an online survey platform to recruit 200 women from a clinical population, a community fair, and the Internet.

**Results:** We recruited 438 women over 29 weeks between September 2017 and March 2018. After consent and eligibility determination, 345 enrolled, 278 started, and 247 completed the survey. Survey initiation and (completion) by recruitment location were 43 (28) from the clinic, 61(60) from a community fair, and 174 (159) from the internet. Among all participants, the mean (SD) age was 27 (6) years, body mass index was 26 kg/m^2^ (7), 79.7% had a college degree or higher, and 14.6% reported a physician diagnosis of PCOS. Race/ethnic distribution was 64.7% White, 11.8% Black, 7.7% Hispanic; and 5.9% Asian; 9.9% reported more than one race/ethnicity. The highest enrollment of Black race/ethnicity was in clinic (40.5%) compared to 1.6% in the community fair, and 8.3% using the internet. Survey completion rates were highest among those recruited from the internet (91.4%) and community fairs (98.4%), compared to in-clinic (65.1%).

**Conclusion:** Multimodal recruitment achieved target recruitment in a short time period, and established a racially diverse cohort to study ovulation and menstruation health. There was greater enrollment and completion rates among those recruited via the internet and community-fair.

**Key Message:** The Ovulation and Menstruation Health Pilot Study:

- Designed to determine the population prevalence of PCOS using a survey instrument and pictorial tool to ascertain menstrual cycle characteristics and androgen excess, and serve as a platform for a future longitudinal cohort study.
- Enrolled participants from diverse backgrounds using an online adaptable platform for multimodal recruitment.
- Mode of recruitment was associated with race/ethnic diversity and completion of survey.

## Introduction

Polycystic ovary syndrome (PCOS), initially described in 1935 [1], is now considered the most common endocrine disorder in reproductive aged women affecting 10% - 15% of these women [2]. Prevalence estimates range from as low as 8% [3] to as high as 26% [4], and depend on the definition used and population studied. The disorder is characterized by clinical or biochemical androgen excess, menstrual irregularity, and the presence of polycystic ovarian morphology on ultrasound visualization [5]. Clinical androgen excess typically presents as acne, hirsutism, or androgenic alopecia [6-8]. Women with PCOS may also experience infertility, insulin resistance and obesity [9-12].

Existing research on PCOS is typically conducted in either small clinical cohorts or larger epidemiologic studies. The latter have variable ascertainment of the disease or disease features which may increase misclassification of disease state and bias estimates of risk [13]. Preliminary studies on PCOS in existing population-based cohorts such as the Nurses’ Health Study 2 (NHS2), the Framingham Heart Study (FHS) and the Cape Cod Health Study (CCHS) are limited by poor correlation of PCOS self-reports compared to medical records (NHS2) [14], too few reported cases of menstrual irregularity (FHS) [15], and inadequate determination of phenotype (CCHS) [16]. Inaccurate classification using single yes/no questions to ascertain those with PCOS may bias estimates of disease.

Existing epidemiologic cohorts are also limited with respect to race and ethnic diversity, as more than 90% of participants were White in these three cohorts [16-18], thereby limiting the ability to detect differences in prevalence and etiological associations across racial and ethnic groups. Racial and ethnic-specific differences in androgen excess and metabolic syndrome symptoms are well established but the reasons for this are not well-understood [19-21].

The Ovulation and Menstruation Health (OM) Pilot Study was conducted to (1) determine the feasibility of enrolling participants from diverse backgrounds using varied recruitment modalities, and (2) understand how survey completion status differed by participant characteristics.

## Materials and Methods

The OM study website consists of a short animated educational recruitment video, an online consent form, screening questions, and a survey instrument. The animated video was designed to appeal to a diverse audience, as its illustrations included women of all races and body types. The goal of the OM Pilot Study was to recruit at least 200 reproductive-aged women over a one-year period. Those who were menstruating or had the capacity to menstruate were eligible (trans-male and other). Participants who were less than 18 or older than 45 years of age, identified as male, pregnant at the time of the survey, had a hysterectomy/oophorectomy, amenorrhea due to radiation or chemotherapy, or were unwilling or unable to provide an email address were deemed ineligible. The survey instrument was publicly accessible from August 21, 2017 to February 26, 2018 on the study website and was used in each recruitment modality. The Boston Medical Center and Boston University Medical Campus Institutional Review Boards approved the study (IRB: H-35075).

### Incentive

The first two hundred participants who completed the survey were entered into a lottery for a $200 gift card. All of those approached were offered free earphones.

### Recruitment

The multimodal recruitment locations were: in-clinic, a Boston-based community event, and the internet. The recruitment approach was adapted to meet the needs of each recruitment location.

### In-Clinic

The in-clinic recruitment occurred from September to October 2017. Approximately 2000 informational recruitment letters including the study website were sent during this recruitment period to patients with an upcoming visit to the Department of Obstetrics and Gynecology (OB/GYN) at Boston Medical Center (BMC). For 20 days, research assistants (RAs) showed the study website and the promotional video via an electronic tablet to interested persons at two OB/GYN waiting rooms.

### A Community Event

A recruitment table was set up at the Boston Women’s Market in the Jamaica Plain neighborhood of Boston, MA on September 17, 2017. This community event was an opportunity for those who identify as female or supporters of women’s causes to sell products from their small businesses.

### Internet

Internet-based recruitment methods included sending out email communications, creation of study social media engagement accounts, and sharing of recruitment materials on individual social networks of the study staff. The study website and social media pages were discoverable on any internet-search. BUMC wide electronic-communications, which included a brief description of the study, eligibility requirements, incentive, and study contact information, began in September, 2017 and ran for the duration of the enrollment period. The study website link was shared on personal networks of study personnel (RM, MN, AW) with the BU Masters of Medical Science Class of 2018 Facebook (FB) group and the Bates Feminist Collective FB in September, 2017 respectively.

Physical flyers and business cards (paper materials) were posted around the BUMC campus for the duration of the study, and at Boston Skin Solutions, a center that treats excess facial and body hair on September 27, 2017.

### Informed Consent, Screening, and Survey Initiation

The online consent form included questions on a participant’s interest in follow-up surveys, other studies, and contributing biospecimens. Once consented, the individual was presented with an eligibility screener. Eligible participants recruited in-clinic were able to start the survey in the waiting room. For eligible participants from the community event individuals and the internet, a unique link to the survey was emailed to the participant’s email address. For all enrolled participants, up to three reminders were sent on consecutive days to those who did not complete the survey.

### Survey Description

The OM Pilot Study survey had a total of 10 sections, with 219 questions; however, most respondents had fewer questions due to skip patterns. The <u>About You</u> section included questions on current residence and whether they ever received care at BMC. The <u>Demographics</u> section included questions on race and ethnicity, education, birth country, and income. The <u>Anthropometrics</u> section captured data regarding height, weight, and body shape using a pictorial tool. The <u>Menstrual Cycle</u> section asked about menarche, regularity of cycles, and cycle tracking. The <u>Hormone Usage</u> section obtained data on hormonal contraceptive usage, history and reasons for hormone usage. The <u>Health and Body</u> section included questions for self-reported androgen excess using a pictorial tool for androgenic alopecia and hirsutism based on the modified Ferriman-Gallwey scale [22]; PCOS status (questions specific to PCOS diagnosis, symptoms for diagnosis, and medication and supplement use, and family history of PCOS); reproductive health (history of infertility, uterine fibroids, endometriosis, and premature ovarian failure); general health (diagnosed medical conditions such as hypertension, diabetes, and other chronic conditions and their treatment); diet and lifestyle (limited questions about alcoholic, non-alcoholic beverage consumption, and smoking habits); obstetric history (ever pregnant, number of prior pregnancies, and each pregnancy outcome, including stillbirths and miscarriages, and live births). For every livebirth, an additional set of questions was asked regarding gestational length and birth outcomes such as birth weight. Study data was recorded and stored on encrypted servers at the Boston University School of Public Health Biostatistics & Epidemiology Data Analytics Center (BEDAC). The OM Pilot Study survey can be found on the Harvard Dataverse [23].

### Data Analysis

Participant characteristics were evaluated by recruitment location and survey completion. Those who started the survey via the internet included those who input the link into their web-browser after learning of the study from any online or printed recruitment materials and completed the online consent and screener. Survey initiation was defined as those who started the “<u>About You”</u> section. Survey completion was defined as having completed the first question of the “<u>Pregnancy and Birth History</u>” upon entering the total number of pregnancies experienced. Those who entered zero pregnancies had no further questions to complete. Those who entered one or more had additional pregnancy related questions to complete. Both nulliparous and multiparous were defined as completing the survey upon entering the total number of pregnancies they experienced. To assess the geographical catchment area associated with the survey, participants’ reported home-state were mapped.

## Results

### Recruitment and Enrollment

Recruitment exceeded our target of 200 women in eight weeks. During the six-month recruitment window, 438 women began the consent process, 384 were assessed for eligibility and 345 women screened-in (88.7% of those assessed for eligibility). Of the 345 participants who consented and screened-in, 278 women started the survey. Among those who started the survey, 66 were recruited in-clinic, 61 from the community event, and 174 from the internet. Figure 1 displays the study enrollment flow chart. Spikes in cumulative enrollment coincided with the active in-clinic recruitment and the community event (Figure 2). Out of the 384 participants who consented and completed the screener, only 39 (10.2%) were excluded. Of the individuals who were excluded, 18 were no longer menstruating, 16 were unwilling or unable to provide email addresses, and five were over the age of 45. Among those who initiated the survey, agreement for re-contact was high for those who responded to re-contact questions, 95.7% consented to be re-contacted for additional information, 80.9% for biological samples, 96.0% for a follow-up survey, and 79.1% for a different research study.

**Figure 1.**
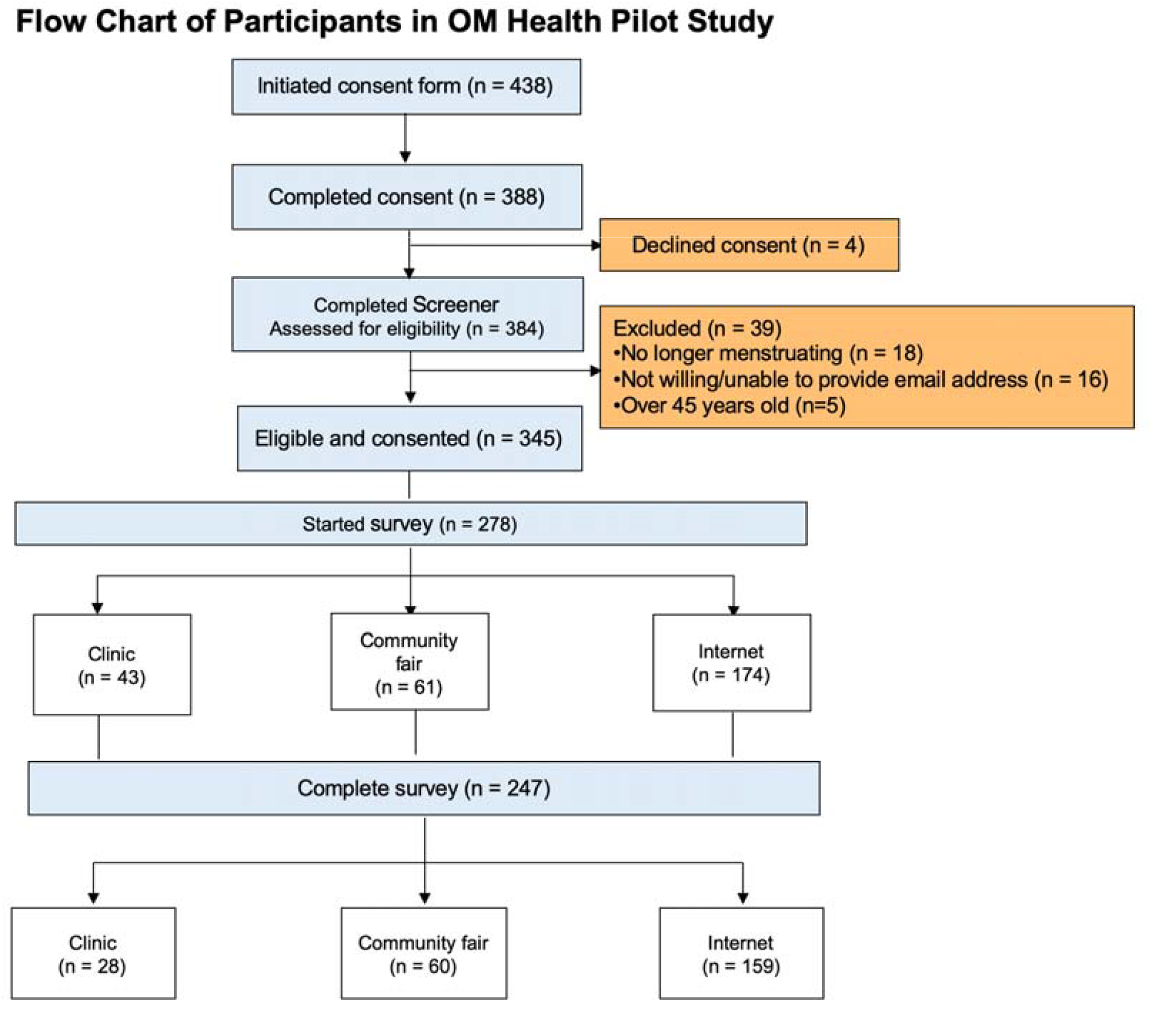
Flow Chart of Participants Enrolled in the OM Health Pilot Study.

**Figure 2.**
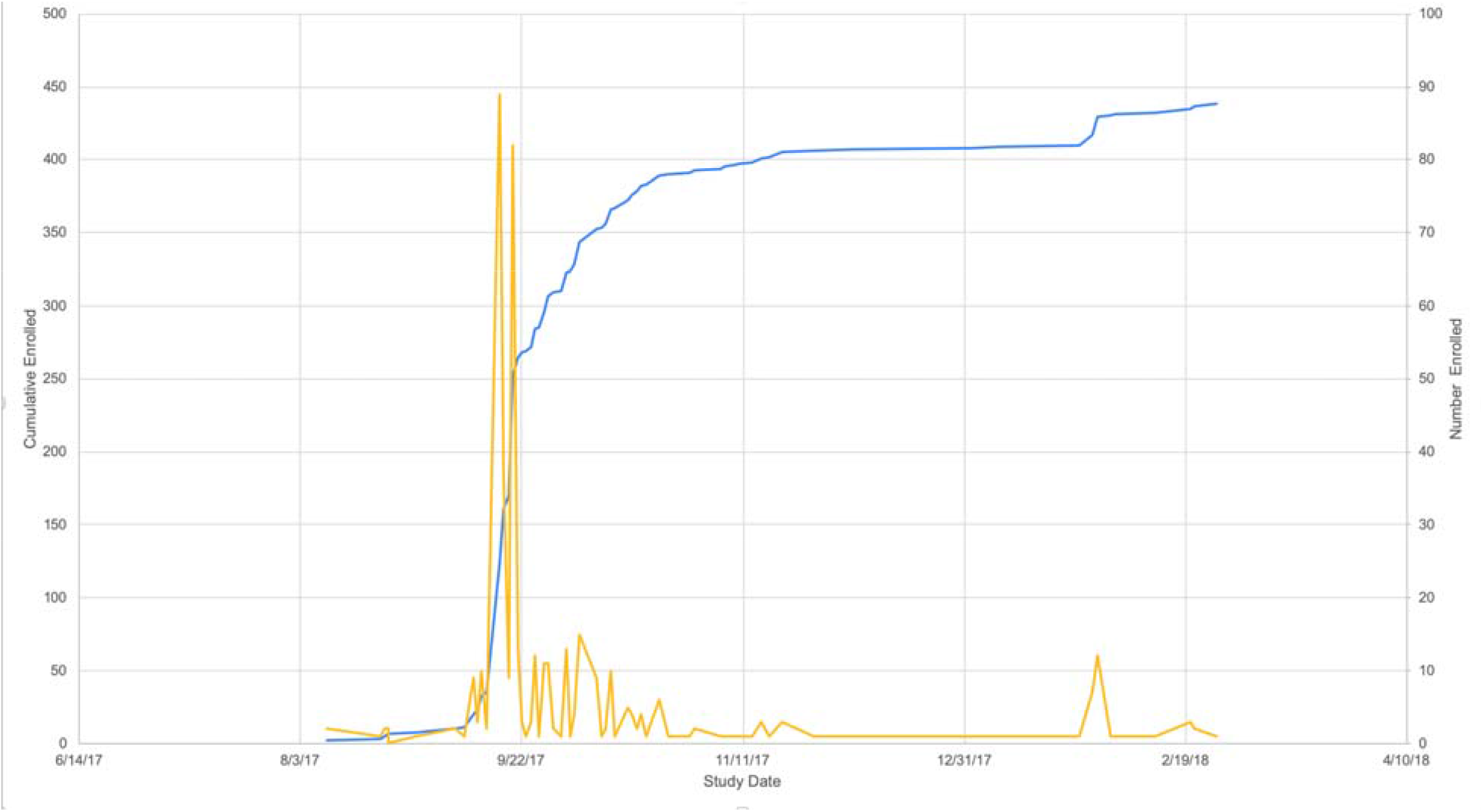
Ovulation and Menstruation Health Pilot Study Enrollment Graph.

### Participant characteristics

Among the 278 women who started the survey, the average age (SD) was 27.3 ± 5.98 years, with the majority (82.1%) born in the United States (Table 1). Participants were White (64.7%), Black/African American (11.8%), Latina/Hispanic (7.7%), and mixed race (9.9%). Two individuals identified as other (non-binary or gender queer). While the majority had a four-year college degree or more (79.7%), 7.0% had a high school degree or less education. Participants were distributed across all income categories, with 21.3% in the less than $25,000 category. The mean (SD) BMI was 26.1 kg/m^2^(6.60), with 15.4% in the overweight category. Of the 278 women who started the survey, 13.1% had smoked at least 100 cigarettes over their lifetime. The reported prevalence of doctor-diagnosed PCOS was 14.6%. Of the 278 women who started the survey, 82.3% had ever used hormonal contraception. The geographic distribution of the cohort included 20 states, as shown in Figure 3. The majority of the participants were recruited from Massachusetts (n=173) and Missouri (n=50).

**Table 1.**
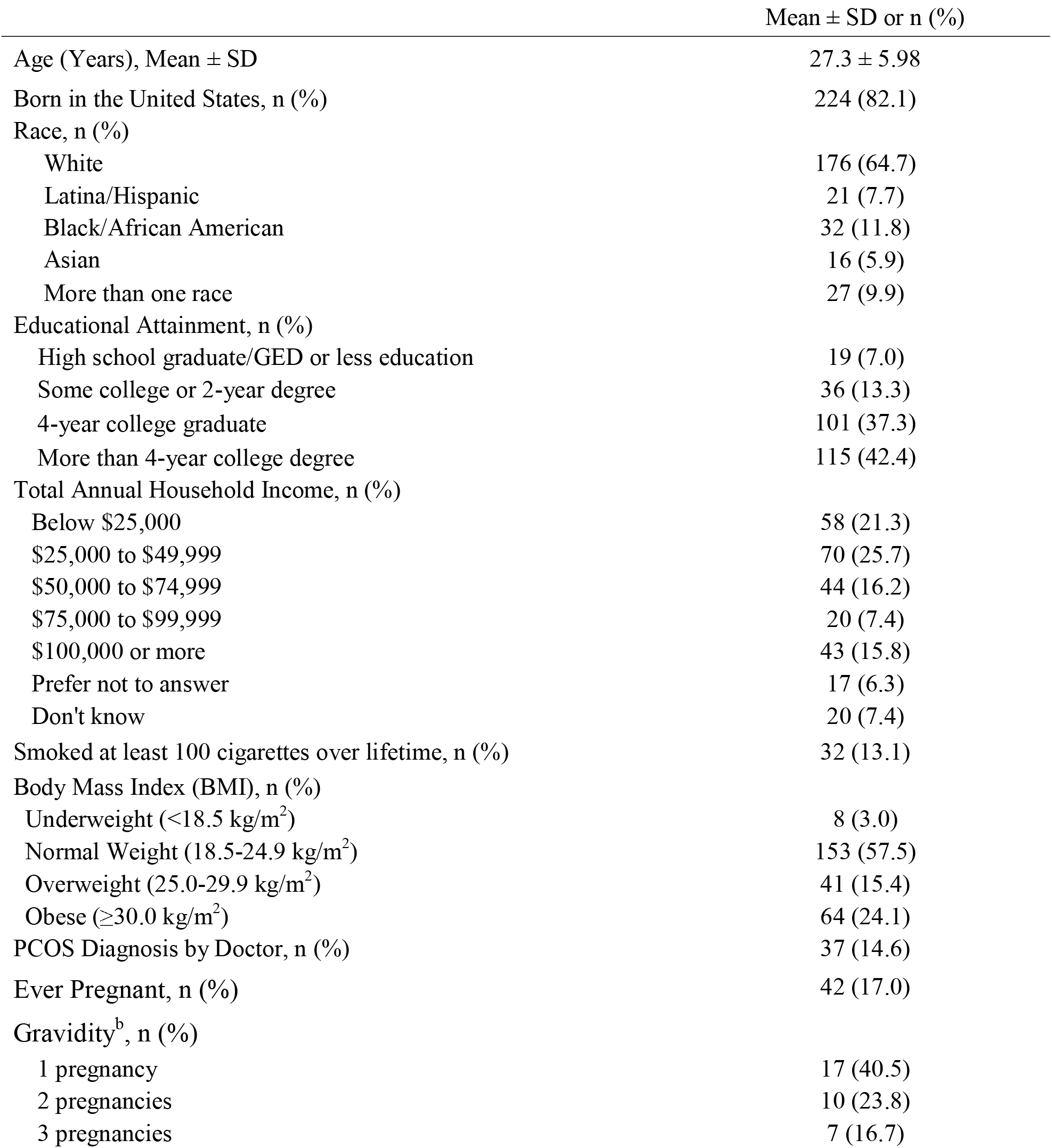
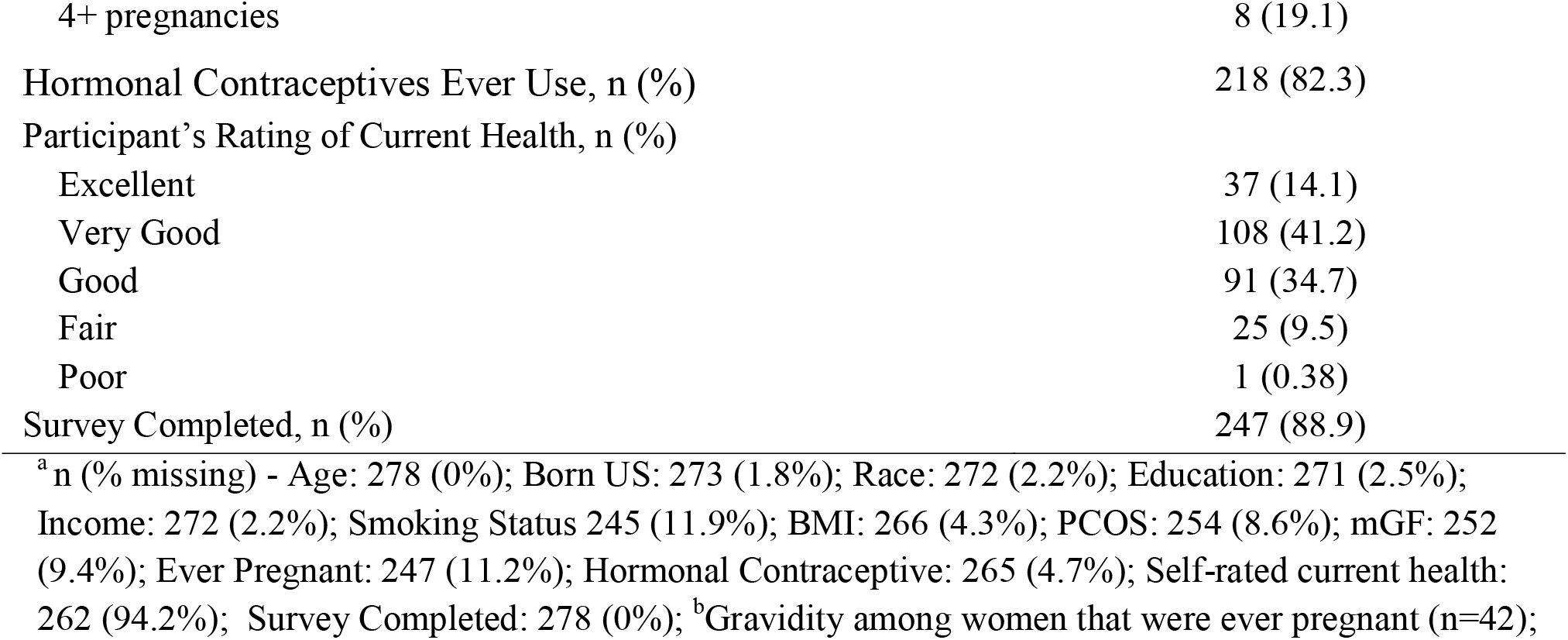
Demographics in the Ovulation and Menstruation Study (OM) Pilot Study (n=278)^a^.

**Figure 3.**
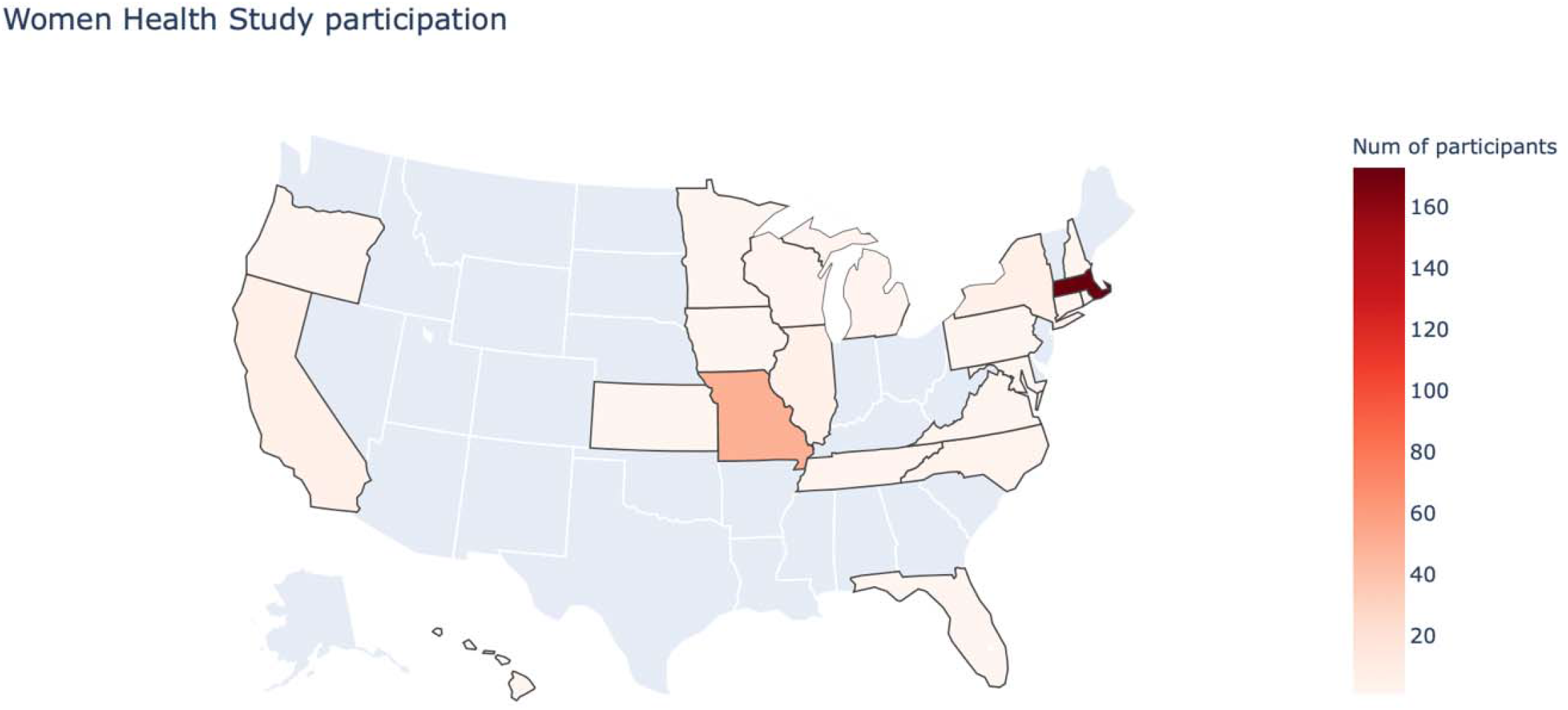
OM Study Participant Population Map.

### Population Demographic Characteristics by Recruitment Location

Table 2 shows participant demographics by recruitment location. Differences were found in age, birth country, race, educational attainment, BMI, and physician-diagnosed PCOS, by location of recruitment. Women recruited in-clinic tended to be older (mean ± SD: 34.0 ± 6.71 years), born outside the US (61.9%), and less likely to have higher educational attainment (19.1% were High school graduate/GED or less education) compared to women recruited at the community fair and via the internet. Women recruited in-clinic were also more likely to be obese (47.4%), have had a physician diagnosis of PCOS (36.4%), and less likely to have ever used oral contraceptives (62.2%) compared to women recruited at the community fair and via the internet. Those from the community fair and internet were more likely to be born in the US (95.1% and 88.2%, respectively), White (77.1% and 71.6%), and have greater than a four-year college degree (39.3% and 49.4%), respectively, than those recruited from the clinic. Women recruited from outside the clinic were younger (community fair: mean 23.0 ± 4.03, internet: mean 25.0 ± 5.36 years), leaner BMI (community fair: 23.3 kg/m^2^ ± 4.79, internet: 23.5 kg/m^2^ ± 6.95), and had a lower prevalence of physician diagnosed PCOS (community fair: 4.9%, internet: 13.8%). Of note, 46.4% of in-clinic-recruited population had been pregnant compared to 3.3% and 17.0% for the community fair and internet participants, respectively. Lastly, income and smoking status did not differ in a meaningful way by recruitment location.

**Table 2.** Demographic Characteristics in the Ovulation and Menstruation Study (OM) Pilot, by Recruitment Location (n=278)

|  | *Clinic*<br>*n=43<sup>a</sup>* | *Community Fair*<br>*n=61<sup>b</sup>* | *Internet*<br>*n=174<sup>c</sup>* |
| --- | --- | --- | --- |
| Age (Years), Median ± SD | 34.0 ± 6.71 | 23.0 ± 4.03 | 25.0 ± 5.36 |
| Born in the United States, n (%) | 16 (38.1) | 58 (95.1) | 150 (88.2) |
| Race, n (%) |  |  |  |
| White | 8 (19.1) | 47 (77.1) | 121 (71.6) |
| Latina/Hispanic | 12 (28.6) | 2 (3.3) | 7 (4.1) |
| Black/African American | 17 (40.5) | 1 (1.6) | 14 (8.3) |
| Asian | 3 (7.1) | 1 (1.6) | 12 (7.1) |
| More than one race | 2 (4.8) | 10 (16.4) | 15 (8.9) |
| Educational Attainment, n (%) |  |  |  |
| High school graduate/GED or less education | 8 (19.1) | 6 (9.8) | 5 (3.0) |
| Some college or 2-year degree | 14 (33.3) | 8 (13.1) | 14 (8.3) |
| 4-year college graduate | 12 (28.6) | 23 (37.7) | 66 (39.3) |
| More than 4-year college degree | 8 (19.1) | 24 (39.3) | 83 (49.4) |
| Total Annual Household Income, n (%) |  |  |  |
| Below \$25,000 | 4 (9.5) | 13 (21.3) | 41 (24.3) |
| \$25,000 to \$49,999 | 12 (28.6) | 20 (32.8) | 38 (22.5) |
| \$50,000 to \$74,999 | 11 (26.2) | 9 (14.8) | 24 (14.2) |
| \$75,000 to \$99,999 | 1 (2.4) | 3 (4.9) | 16 (9.5) |
| \$100,000 or more | 5 (11.9) | 6 (9.8) | 32 (18.9) |
| Prefer not to answer | 8 (19.1) | 2 (3.3) | 7 (4.1) |
| Don't know | 1 (2.4) | 8 (13.1) | 11 (6.5) |
| Smoked at least 100 cigarettes over lifetime, n (%) | 5 (17.9) | 5 (8.3) | 22 (14.0) |
| Body Mass Index (BMI), n (%) |  |  |  |
| Normal Weight or Underweight ($\leq 25$ kg/m<sup>2</sup>) | 14 (36.8) | 40 (65.6) | 107 (64.1) |
| Overweight (25-30 kg/m<sup>2</sup>) | 6 (15.8) | 11 (18.0) | 24 (14.4) |
| Obese ($\geq 30$ kg/m<sup>2</sup>) | 18 (47.4) | 10 (16.4) | 36 (21.6) |
| PCOS Diagnosis by Doctor, n (%) | 12 (36.4) | 3 (4.9) | 22 (13.8) |
| Ever Pregnant, n (%) | 13 (46.4) | 2 (3.3) | 27 (17.0) |
| Gravidity<sup>d</sup>, n (%) |  |  |  |
| 1 pregnancy | 5 (38.5) | 1 (50.0) | 11 (40.7) |
| 2 or more pregnancies | 8 (61.5) | 1 (50.0) | 16 (59.3) |
| Hormonal Contraceptives Ever Use, n (%) | 23 (62.2) | 54 (88.5) | 141 (84.4) |
| Participant's Rating of Current Health, n (%) |  |  |  |
| Excellent | 6 (16.2) | 8 (13.1) | 23 (14.0) |
| Very Good | 7 (18.9) | 29 (47.5) | 72 (43.9) |
| Good | 18 (48.7) | 21 (34.4) | 52 (31.7) |
| Fair to Poor | 6 (16.2) | 3 (4.9) | 17 (10.4) |
| Survey Completed, n (%) | 28 (65.1) | 60 (98.4) | 159 (91.4) |

<sup>a</sup> n (% Missing) - Age: 43 (0%); Born US: 42 (2.3%); Race: 42 (2.3%); Education: 42 (2.3%); Income: 42 (2.3%); Smoking Status:
28 (34.9%); BMI:38 (11.6%); PCOS: 33 (23.3%); Ever Pregnant: 28 (34.9%); Hormonal Contraceptive: 37
(13.9%); Self-rated current health: 37 (14.0%); Survey Completed: 43 (0%).
<sup>b</sup> n (% Missing) - Age: 61 (0%); Born US: 61 (0%); Race: 61 (0%); Education: 61 (0%); Income: 61 (0%); Smoking Status 60 (1.6%);
BMI:61(1.6%); PCOS: 61 (0%); Ever Pregnant: 60 (1.6%); Hormonal Contraceptive: 61 (0%); Self-rated current
health: 61 (0%); Survey Completed: 61 (0%).
<sup>c</sup> n - Age: 174 (0%); Born US: 170 (2.3%); Race: 169 (2.9%); Education: 168 (3.5%); Income: 169 (2.9%); Smoking Status 157
(9.8%); BMI:167 (4.0%); PCOS: 160 (8.1%); Ever Pregnant: 159 (8.6%); Hormonal Contraceptive: 167 (4.0%); 164
(5.7%); Survey Completed: 174 (0%).
<sup>d</sup>Gravidity among women that were ever pregnant (n=42).

### Survey Completion

Among the 278 women who started the survey, 247 (88.8%) completed it. The median time to completion was 12.6 minutes (range: 1.03 minute -92 days). Seven participants took more than 24 hours to complete the survey. Women completing the survey were more likely to be US born (85.7% vs. 51.9%), White (67.4% vs. 40.7%) and have 4 years of college education or more (81.9% vs. 59.2%) compared to those who did not complete the survey (Table 3). We also found that the women who did not complete the survey were older age and had higher annual household income compared to those who completed the survey. Among the 31 women who did not complete the survey, survey drop off seemed equally distributed across section categories, show in Figure 4.

**Table 3.** Demographic Characteristics in the Ovulation and Menstruation Study (OM) Pilot, By Survey Completion Status (n=278)

|  | *Complete*<br>*(n=247)<sup>a</sup>* | *Incomplete*<br>*(n=31)<sup>b</sup>* |
| --- | --- | --- |
| Age (Years), Median ± SD | 25.0 ± 5.68 | 29.0 ± 7.37 |
| Born in the United States, n (%) | 210 (85.7) | 14 (51.9) |
| Race, n (%) |  |  |
| White | 165 (67.4) | 11 (40.7) |
| Latina/Hispanic | 15 (6.1) | 6 (22.2) |
| Black/African American | 25 (10.2) | 7 (25.9) |
| Asian | 15 (6.1) | 1 (3.7) |
| More than one race | 25 (10.2) | 2 (7.4) |
| Educational Attainment, n (%) |  |  |
| High school graduate/GED or less education | 14 (5.7) | 5 (18.5) |
| Some college or 2-year degree | 30 (12.3) | 6 (22.2) |
| 4-year college graduate | 94 (38.5) | 7 (25.9) |
| More than 4-year college degree | 106 (43.4) | 9 (33.3) |
| Total Annual Household Income, n (%) |  |  |
| Below \$25,000 | 54 (22.0) | 4 (14.8) |
| \$25,000 to \$49,999 | 63 (25.7) | 7 (25.9) |
| \$50,000 to \$74,999 | 42 (17.1) | 2 (7.4) |
| \$75,000 to \$99,999 | 19 (7.8) | 1 (3.7) |
| \$100,000 or more | 39 (15.9) | 4 (14.8) |
| Prefer not to answer | 11 (4.5) | 6 (22.2) |
| Don't know | 17 (6.9) | 3 (11.1) |

<sup>a</sup>n (% Missing) - Born US: 246 (0.40%); Race: 245 (0.81%); Education: 244 (1.21%); Income: 245 (0.81%).
<sup>b</sup>n (% Missing) - Born US: 27 (12.9%); Race: 27 (12.9%); Education: 27 (12.9%); Income: 27 (12.9%).

**Figure 4.**
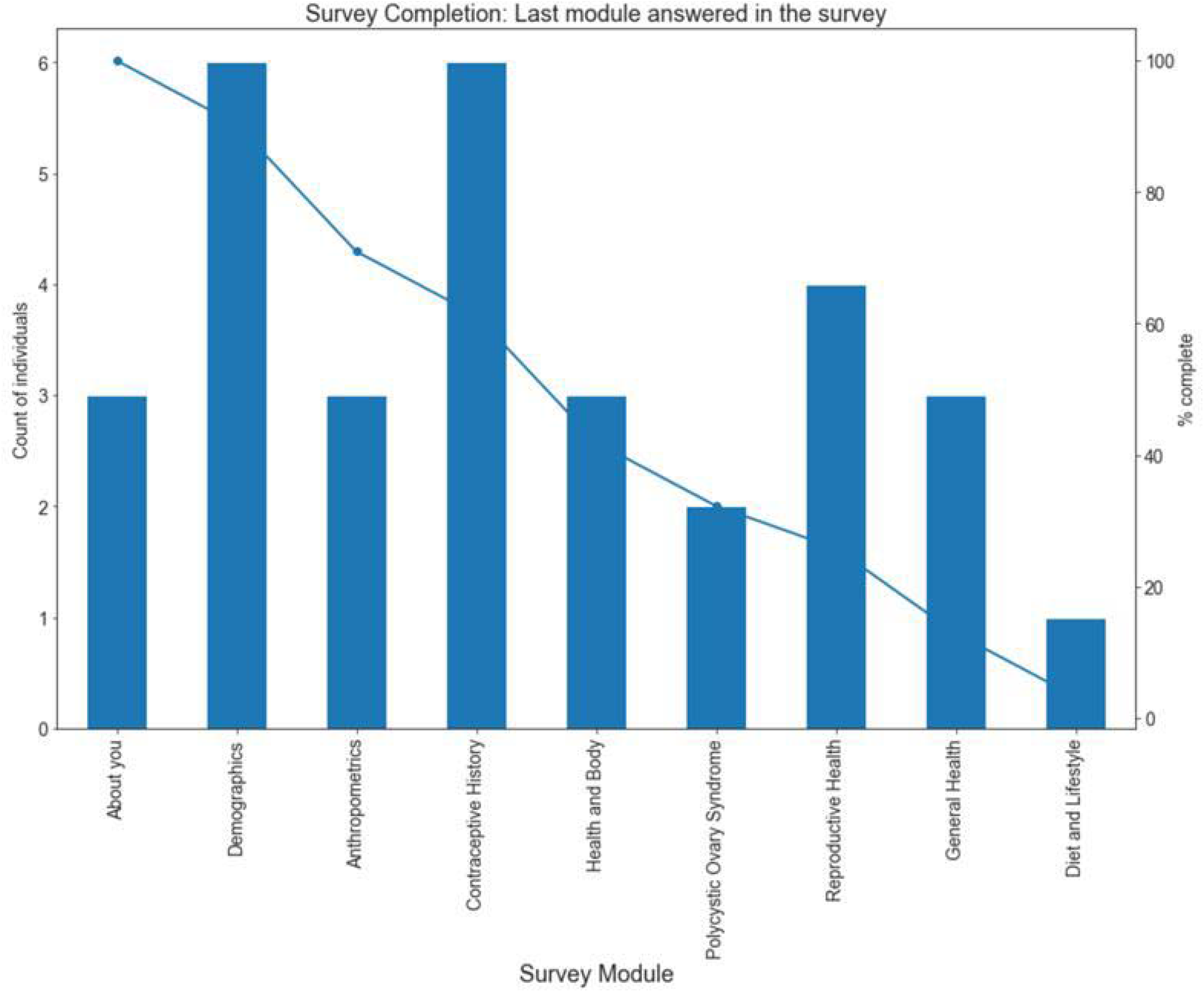
Survey Completion Bar Graph with Count Line Overlay.

## Discussion

To the best of our knowledge, this is the first study to determine the feasibility of a multimodal recruitment approach for a study of ovulation and menstruation health, comparing in-clinic, community-based, and internet-based recruitment activities. Among the recruitment modalities, the majority of women were recruited via the internet after encountering advertising materials informing them of the study website. The race/ethnicity of the cohort was notably similar to the US population [24].

The participant characteristics did differ by recruitment location, suggesting that multimodal recruitment is a feasible solution to increase the variation in participant characteristics including race/ethnicity and health characteristics, such as BMI, oral contraceptive use, and physician diagnosed PCOS. These different characteristics may reflect differences in the source population. For example, approximately 72% of the patients at BMC come from an underserved population [25] whereas the community event was held in Jamaica Plain, a neighborhood that has a 53.6% White population, and a median household annual income of $55,861 [26]. While use of the internet is reported at 92% of Whites, 86% of Hispanic, and 85% of Blacks in the U.S. [27], those accessing the study via the internet were recruited through flyers, email communications, and sharing with personal social media networks. The geographic distribution of the participants was notable for the majority in Massachusetts, with second highest participation in Missouri, corresponding to the personal network of one research assistant.

Similarly, the All of Us Study, was also successful in recruiting racially and ethnically diverse participants using a combination of 340 recruitment sites and digital campaigns [28]. Among the All of Us participants, 51% of their core participants were considered “nonwhite” whereas participants in traditional reproductive health cohorts such as Boston University Pregnancy Study Online cohort were 83% Caucasian [29], 89% White in the Nurses’ Health Study 2, and 100% in the original Framingham Heart Study. However, racial diversity steadily increased in subsequent Framingham-related studies such as the Omni Cohort which was comprised of 28% African American, 42% Hispanic American, and 24% Asian American participants [30].

The completion rate among those who started our survey (88.8%) was high. This completion rate is comparable to the Pregnancy Study Online (PRESTO) with 72% of enrolled participants completing the baseline survey [29] and 85% for the most recent Framingham Heart Study participants using an electronic survey [31]. It is possible that our survey was shorter than those studies and as such had a slightly higher completion rate. Furthermore, the OM survey was designed for an 8^th^ grade reading level and underwent cognitive and usability testing to facilitate question comprehension, potentially supporting a pleasant participant experience [32]. While participants could skip any questions, the rate of missingness was low. Birthplace outside of the US was a key variable noted among the small proportion of women who did not complete the survey.

This study has several strengths. We achieved target recruitment in half the expected time, which may have been due to an appealing online study platform that included an educational cartoon featuring women from multiple backgrounds or the monetary incentive. Use of a cognitively tested survey designed for an 8^th^ grade reading level may also have facilitated the high completion rate. The broad inclusion criteria also allowed for those with and without PCOS to be included. Most importantly, the multimodal recruitment approach enabled us to recruit a more diverse cohort, which is reflective of studies using a similar multimodal recruitment strategy for other health conditions [33].

However, the present study has some key limitations. While the racial and ethnic diversity was improved compared to existing large epidemiologic cohorts, the proportion of Latina/Hispanic women was slightly lower than the US population, possible due to language access issues. We recently translated the OM Platform for availability in Spanish language to test in future studies. Also, completion rate by those born outside the US may be improved by having the survey in the primary language of the participant. In the future, using paid advertisements may not only lead to high click-through rates to the study website, but may also provide for a unique opportunity to engage with specific targeted demographics depending on advertisement dissemination. For example, NHS3 sent recruitment postcards to minority-dense zip codes doubling the enrollment rate of African American and Hispanic women [17].

Furthermore, we considered whether bias in multimodal recruitment exists. We do not believe that multimodal recruitment increased the likelihood of selection bias since enrollment would need to be related to both the exposure and outcome under study for such a bias to occur [34-37]. The multimodal recruitment strategy increases the generalizability of the study and increases its ability to examine effect measure modification by racial and ethnic group by ensuring sufficient numbers in each category. Because differences exist in racial/ethnic-specific risks for a variety of health outcomes potentially due to structural racism and other inequities [38], a multi-ethnic cohort is needed to assess health outcomes subsequent to PCOS diagnosis and to determine ideal windows for risk-reducing interventions.

## Conclusion

Multimodal recruitment was feasible and established a more racially and ethnically diverse cohort to study ovulation and menstruation health, as compared to prior studies. The sampling from the three different recruitment locations demonstrates variability in racial, ethnic and other demographic and health related features.

## Data Availability

The OM Pilot Study survey can be found on the Harvard Dataverse.

https://dataverse.harvard.edu/dataset.xhtml?persistentId=doi:10.7910/DVN/XACYQA

## Acknowledgements

We would like to acknowledge the support of the Department of OB/GYN at Boston University Medical Campus.

